# Deep Learning for Automated Meningioma Segmentation: Toward Clinical Integration and Workflow Efficiency

**DOI:** 10.64898/2026.05.12.26352585

**Authors:** Ebru Fenney, Laxmi Muralidharan, James K Ruffle, Anand Pandit, Mirabel Millip, Ahmed Hammam, Thomas Brookes, Farrah Jabeen, Jordan Colman, Omran Sarwani, Komeil Alattar, Evgenia Efthymiou, Neha Kallam, Juveria Siddiqui, Hani J Marcus, Parashkev Nachev, Harpreet Hyare

**Affiliations:** Department of Translational Neuroscience and Stroke, UCL Queen Square Institute of Neurology, London, United Kingdom; Lysholm Department of Neuroradiology, National Hospital for Neurology and Neurosurgery, London, United Kingdom; Department of Neurosurgery, National Hospital for Neurology and Neurosurgery, London, United Kingdom

**Keywords:** meningioma, segmentation

## Abstract

**Key Results:**

1. In five-fold cross-validation (1000 cases, six institutions), the model achieved mean Dice similarity coefficients of 0.939 for enhancing tumor, 0.937 for tumor core, and 0.921 for whole tumor, with tumor core volumes strongly correlated with reference volumes (r = 0.995).
2. In external validation (310 cases, single institution), mean tumor core Dice was 0.872 despite heterogeneous MRI protocols and incomplete sequences; tumor core volume correlation remained strong (r = 0.971).
3. In a blinded evaluation by 10 radiologists across 510 cases, model segmentations scored higher than reference annotations, with the advantage fourfold larger in real-world clinical data; mean inference time was 1.2 seconds per case.

**Summary Statement:** A fully automated deep learning model achieved high meningioma segmentation accuracy, generalized to heterogeneous clinical imaging in 1.2 seconds, and surpassed reference annotation quality in blinded radiologist evaluation.

**Background:** Meningiomas are the most common primary intracranial tumors in adults, and volumetric assessment increasingly guides surveillance and treatment decisions. Automated segmentation could enable standardized volumetry but requires robust validation.

**Purpose:** To develop a fully automated three-dimensional deep learning model for meningioma segmentation on multiparametric MRI, and to evaluate segmentation accuracy, external generalizability, failure modes, radiologist-rated clinical plausibility, and workflow feasibility.

**Methods:** From 2024 to 2026, this retrospective study trained a custom 3D nnU-Net residual encoder model. Expert segmentations covered enhancing tumor (ET), tumor core (TC), and whole tumor (WT). Dice similarity coefficient (DSC) was the primary metric. External validation used an independent single-institution dataset (n = 310 intracranial cases) with incomplete MRI protocols. Failure modes, model equity, and inference time were assessed. A blinded multi-rater study (10 radiologists; 510 cases) rated TC segmentations using a 0–10 Likert scale, analyzed with linear mixed-effects models.

**Results:** Model training used the BraTS Meningioma 2023 dataset (n = 1000; mean age 60.2 ± 14.5; 705 female). In cross-validation, mean DSC was 0.939 for ET, 0.937 for TC, and 0.921 for WT. In external validation, mean DSC was 0.872 for TC and 0.842 for WT, despite heterogeneous protocols and incomplete sequences. Predicted TC volumes correlated strongly with reference volumes in cross-validation (r = 0.995) and external validation (r = 0.971). Most common failure modes were skull base and intraosseous tumors with performance equitable across demographic subgroups. Mean inference time was 1.2 seconds. In blinded evaluation (1120 ratings), model segmentations received higher scores than reference annotations (+0.32 BraTS; +1.38 external validation).

**Conclusion:** A fully automated deep-learning model achieved high meningioma segmentation accuracy across multi-institutional training data and external clinical imaging. In a blinded study, model segmentation quality exceeded reference annotations, and 1.2-second inference supported workflow integration. Prospective evaluation is warranted before routine deployment.

## Introduction

Meningiomas are the most common primary intracranial tumor in adults and account for approximately one-third of central nervous system neoplasms (1–3). Most are World Health Organization (WHO) grade 1, although a clinically important minority demonstrate aggressive behavior, including rapid growth, recurrence, or malignant transformation (1,4,5). MRI is central to diagnosis, preoperative planning, radiotherapy targeting, and longitudinal surveillance (4).

Volumetric assessment provides a more accurate representation of tumor burden and is increasingly used in research and specialist clinical practice, particularly for surveillance of incidental meningiomas where growth rate influences management decisions (6,7). Serial volumetric studies have demonstrated heterogeneity in meningioma growth patterns and have supported the use of objective volumetry to guide timing of intervention (6–8).

Despite its clinical relevance, volumetric reporting is not routinely performed in standard radiology workflows. Manual segmentation is time-consuming and requires specialist expertise. It is also subject to interobserver variability, particularly for lesions with complex margins adjacent to dura, venous sinuses, cranial nerves, or bone. Variability is further amplified when delineating associated edema on FLAIR imaging, where boundaries may be indistinct and labeling conventions vary. Together these limit standardization across readers and institutions and reduce feasibility for routine longitudinal volumetric follow-up.

Deep learning–based segmentation has improved performance across a range of neuro-oncologic imaging tasks and has become a dominant approach for automated volumetry (9,10). In meningiomas, convolutional neural network models have demonstrated feasibility for automated segmentation on routine multiparametric MRI and have shown potential for reducing the time burden of manual contouring (11,12,13). However, a key barrier to clinical use is generalizability: many published models are trained using single-institution datasets with relatively homogeneous scanner hardware and acquisition protocols, which may inflate apparent performance and reduce robustness when applied to heterogeneous real-world imaging (14). Additionally, follow-up examinations may include only contrast-enhanced T1-weighted imaging, whereas many segmentation models assume the availability of full multiparametric MRI, causing potential failure in common clinical scenarios.

Evaluation methodology also complicates the interpretation of published performance. Overlap metrics such as Dice similarity coefficient (DSC) are widely used but assume accurate and complete reference annotations (15). In practice, “ground truth” segmentations may contain omissions, inconsistencies, or systematic biases, particularly for edema and for small lesions, causing DSC to underestimate true model performance (15,16). External validation and radiologist-centered evaluation provide more robust testing and assessment of clinical impact.

Therefore, the purpose of this study was to develop and evaluate a custom fully automated meningioma segmentation model using a three-dimensional nnU-Net residual encoder framework and to assess segmentation performance, external generalizability, failure modes, radiologist-rated segmentation quality, and inference time as a measure of workflow feasibility (10).

## Materials and Methods

### Study Design

This retrospective study developed and evaluated a fully automated meningioma segmentation model via five steps: cross-validation, external validation, model equity analysis, inference timing, and a blinded multi-rater study (Figure 1).

**Figure 1:**
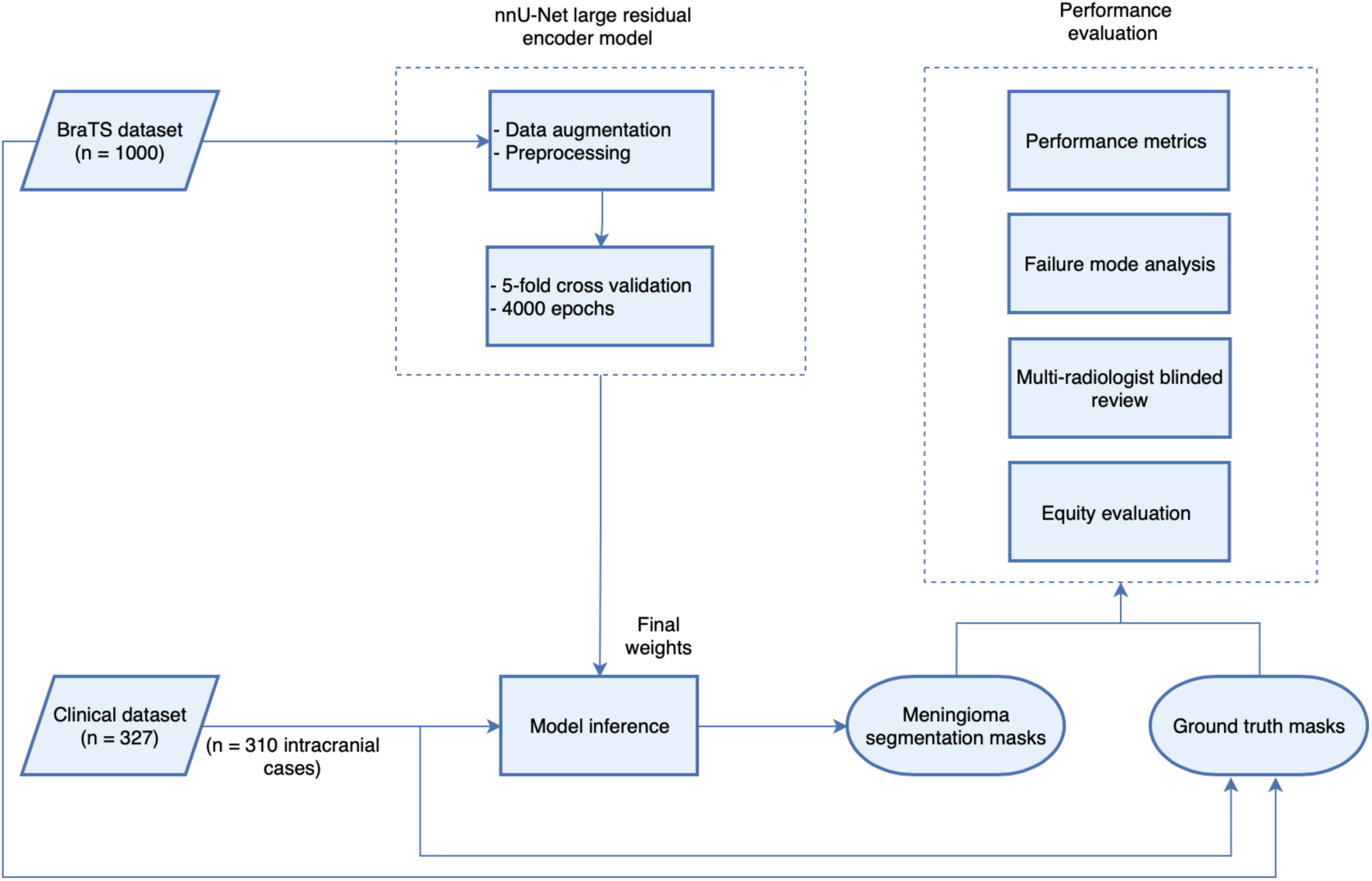
Flowchart of model development. Overview of the model development and validation pipeline. Cross-validation: training was performed on the multi-institutional BraTS Meningioma 2023 dataset (n = 1000) using multiparametric MRI (T1-weighted, contrast-enhanced T1-weighted, T2-weighted, and FLAIR). Preprocessing included resampling to standardized voxel spacing and Z-score intensity normalization; data augmentation incorporated affine, elastic, intensity, and noise transforms alongside channel dropout to improve robustness to missing sequences. A 3D nnU-Net residual encoder was trained with five-fold cross-validation (4000 epochs per fold; Dice–cross entropy loss) to segment the three BraTS label classes (enhancing tumor, non-enhancing core, peritumoral FLAIR hyperintensity), from which the evaluation regions enhancing tumor (ET), tumor core (TC) (= ET + non-enhancing core), and whole tumor (WT) (= TC + peritumoral hyperintensity) were computed.. External validation was performed on an independent single-institution clinical dataset (n = 310 intracranial cases). Performance was assessed using Dice similarity coefficient (DSC) and Hausdorff distance at the 95th percentile (HD95), with additional evaluation of volumetric agreement (Pearson correlation; Bland–Altman analysis), failure mode analysis, model equity (Gini index), inference time, and blinded radiologist quality rating (0–10 Likert scale; 10 raters; 510 cases).

### Cross-validation

#### Data Source and Preprocessing

The model was trained on the BraTS Meningioma 2023 dataset (n = 1000; six institutions; Table 1) (17). Multiparametric MRI included T1-weighted, contrast-enhanced T1-weighted, T2-weighted, and FLAIR sequences; full scanner acquisition parameters for the external validation dataset are provided in Supplementary Table S1. Provided expert-refined labels for enhancing tumor, non-enhancing/necrotic tumor core, and peritumoral FLAIR hyperintensity were combined into the standard BraTS evaluation regions: enhancing tumor (ET), tumor core (TC = ET + non-enhancing core), and whole tumor (WT = TC + peritumoral hyperintensity). All imaging data were provided in de-identified DICOM format by the BraTS consortium; no additional anonymization was performed by the study authors.

**Table 1:** Clinical characteristics of the training (BraTS) and external validation (clinical) datasets.

| Parameter | Training ($n = 1000$) | External Test ($n = 327$) |
| --- | --- | --- |
| Age (y) | 60.2 ± 14.5 | 67.5 ± 13.8 |
| Age availability | 989 (98.9) | 311 (95.1) |
| Sex |  |  |
| Female | 705 (70.5) | 231 (70.6) |
| Male | 285 (28.5) | 80 (24.5) |
| Unavailable | 10 (1.0) | 16 (4.9) |
| WHO grade |  |  |
| Grade 1 | 526 (52.6) | 257 (78.6) |
| Grade 2 | 158 (15.8) | 48 (14.7) |
| Grade 3 | 18 (1.8) | 3 (0.9) |
| Unavailable | 298 (29.8) | 19 (5.8) |

*Note: Unless otherwise specified, data is number of patients, with percentages in parentheses. $n = 327$ prior to exclusion of 17 extracranial cases; 310 used for analysis.*

Preprocessing included resampling to standardized voxel spacing, Z-score intensity normalization, and augmentation (affine, elastic, intensity, and noise augmentation) to increase the heterogeneity of the training data and enhance generalizability of the model.

#### Model Architecture and Training

A self-configuring 3D nnU-Net residual encoder framework, widely validated for biomedical segmentation (10), was implemented. The 3D full resolution configuration was trained using five-fold cross-validation, with each fold running for 4000 epochs. Optimization employed the Dice–cross entropy hybrid loss. Additional channel dropout (p = 0.3 per sequence) was applied to improve robustness to missing MRI inputs. Final predictions were an equal-weight ensemble of all five-fold checkpoints, consistent with the nnU-Net default configuration; no additional hyperparameter tuning was performed after cross-validation.

#### Evaluation Metrics

The primary metric was Dice similarity coefficient (DSC), with 95th-percentile Hausdorff distance (HD95) reported as a secondary metric, both computed per tumor component (ET,TC,WT). The lowest-performing 5% of cases, by tumor core DSC score, during both cross validation and external validation were reviewed by a consultant neuroradiologist to identify failure patterns and assess for annotation limitations. Cases with misleading poor edema class metrics, attributed to inconsistent reference annotations, were identified by a consultant neuroradiologist and excluded from final DSC calculations.

### Sequence Dropout Ablation

Sequence modality dropout ablation experiments were carried out at inference time during cross validation on the full training set (n=1000). For each held-out validation case, inference was run with all 15 non-empty subsets of the four input modalities. Dice score and HD95 were computed per case for each tumor sub-region (ET, TC, WT, ED) and pooled across all cross-validation folds.

### External Validation

#### Dataset

An independent single-institution dataset of 327 meningioma patients was used for external validation (Table 1). The institutional dataset component was approved by the local research ethics committee, with waiver of informed consent where applicable. All imaging data were anonymized in accordance with institutional information governance policy prior to analysis, with patient identifiers removed from DICOM headers. After excluding extracranial tumors, 310 intracranial patients were retained. Imaging completeness varied, including examinations with only contrast-enhanced T1-weighted sequences. Missing modalities were zero-imputed and handled by the channel dropout augmentation applied during training.

Expert-labeled annotations were available for tumor core and edema. Therefore, tumor core and whole tumor DSC and HD95 were calculated, but enhancing tumor performance could not be evaluated. Tumor core volumes were derived from model predictions and reference annotations. For 154 tumors, the edema segment had not been annotated and for these cases, whole tumor analysis could not be performed. Agreement was assessed using Pearson correlation coefficient and Bland–Altman analysis.

### Model Equity Analysis

To evaluate whether the model performed equitably across demographic subgroups of the test set, analysis was performed using Fairboard (18), a platform for comprehensive fairness and statistical analysis of model performance. Variables assessed included all available metadata from the test set including age, sex, and tumor grade, each evaluated against all tumor component DSC values. The Gini index metric was selected to quantify fairness, ranging from 0 to 1, with lower values indicating more equitable distribution of model performance across all samples.

### Inference Performance

Inference time per case was measured on an NVIDIA RTX PRO 5000 GPU to assess feasibility for clinical workflow integration.

### Radiologist Quality Assessment

Ten raters (3 consultant neuroradiologists; 7 radiology registrars) blindly compared model and reference segmentations across 510 cases (all 310 clinical cases and 200 random BraTS cases). Each scored between 55 and 60 cases randomly drawn from both datasets (∼10% overlap among raters). For each case, contrast-enhanced T1-weighted images were displayed with overlaid tumor core segmentations from each source, and raters scored each method on a 0–10 Likert scale blinded to source. Edema overlays were excluded to avoid bias from inconsistent reference annotations.

Scores were analyzed using a linear mixed-effects model (LMM) in R (lme4/lmerTest) with fixed effects for method, dataset, and their interaction, and random intercepts for rater, case, and rater-by-method (full specification in Supplement S1). Inter-rater reliability on overlapped-cases was assessed with an intercept-only model; intraclass correlation coefficient (ICC) values for absolute agreement and consistency were estimated from variance components.

This study was reported in line with the Checklist for Artificial Intelligence in Medical Imaging (CLAIM) (19).

## Results

### Cross-validation Performance

Mean DSC was 0.939 ± 0.125 for enhancing tumor, 0.937 ± 0.131 for tumor core, and 0.921 ± 0.156 for whole tumor (Table 2). Mean HD95 was 4.202 ± 15.136 mm for ET, 4.198 ± 14.968 mm for TC, and 4.949 ± 15.793 mm for WT. Predicted tumor core volumes demonstrated strong correlation with reference volumes in cross-validation dataset (r = 0.995, 95% CI [0.993, 0.996], Figure 2A).

**Figure 2:**
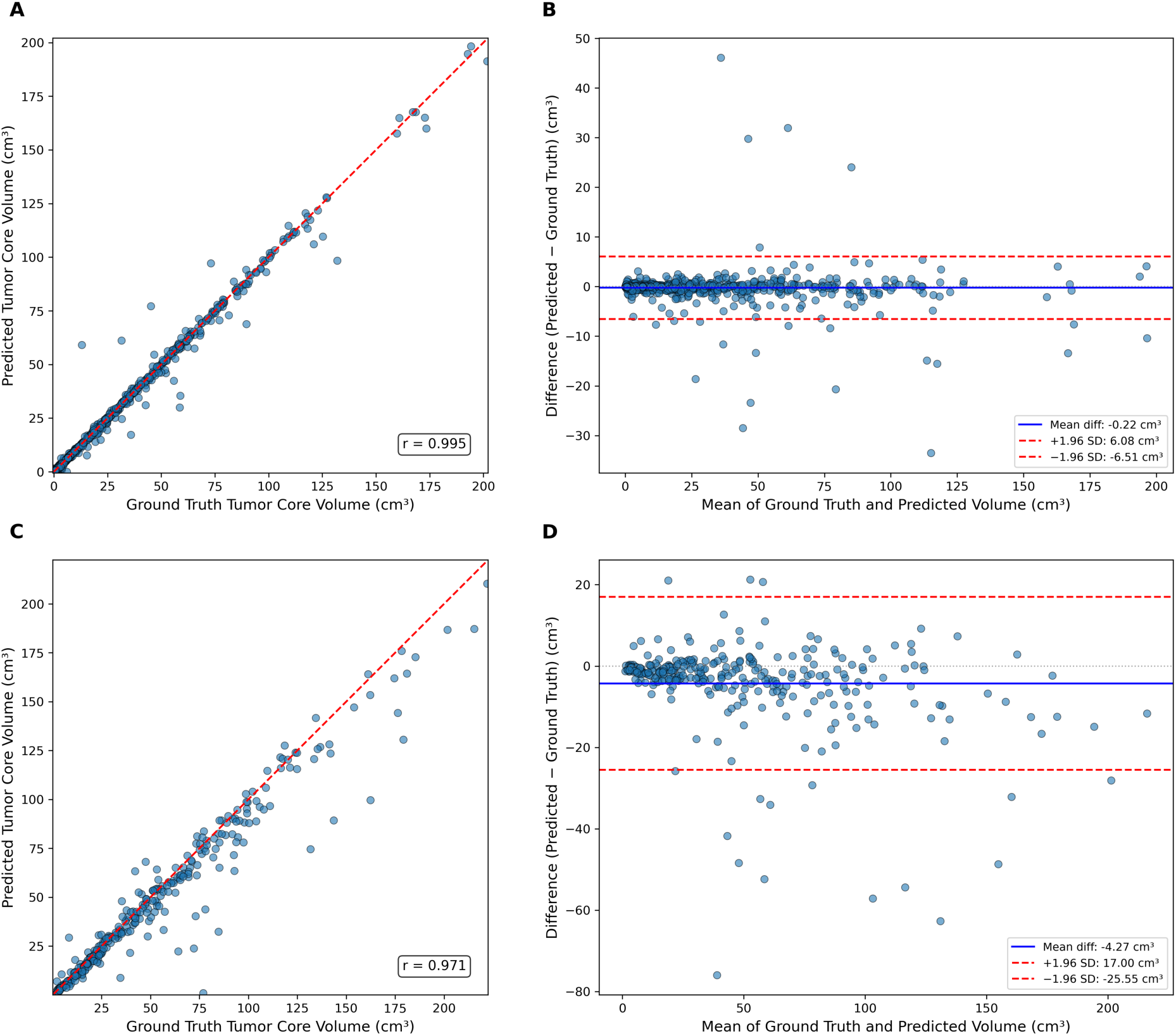
Scatterplot comparing expert-derived tumor volumes and model predictions. Scatterplot comparing expert-derived tumor volumes (x-axis) and model predictions (y-axis). Strong agreement is observed across a wide range of tumor sizes. [A,B] demonstrate cross-validation predicted tumor volume results and [C,D] demonstrate external validation dataset predicted tumor volume results.

**Table 2:** Lesion-wise cross-validation (BraTS dataset) metrics.

| Region | DSC |  | HD95 |  | *n* |
| --- | --- | --- | --- | --- | --- |
| | Mean ± Std | 95% CI | Mean ± Std | 95% CI | |
| ET | $0.939 \pm 0.125$ | [0.931, 0.946] | $4.202 \pm 15.136$ | [3.303, 5.170] | 1000 |
| TC | $0.937 \pm 0.131$ | [0.928, 0.945] | $4.198 \pm 14.968$ | [3.284, 5.148] | 1000 |
| ED (before excl.) | $0.509 \pm 0.448$ | [0.478, 0.539] | $4.633 \pm 11.805$ | [3.810, 5.575] | 1000 |
| ED (after excl.) | $0.822 \pm 0.259$ | [0.799, 0.843] | $4.027 \pm 9.822$ | [3.326, 4.814] | 678 |
| WT (before excl.) | $0.921 \pm 0.142$ | [0.912, 0.930] | $4.651 \pm 15.207$ | [3.724, 5.627] | 1000 |
| WT (after excl.) | $0.921 \pm 0.156$ | [0.909, 0.932] | $4.949 \pm 15.793$ | [3.812, 6.238] | 678 |

*Note: ET = enhancing tumor, TC = tumor core, ED = edema, WT = whole tumor. 322 cases were excluded from final edema and whole tumor evaluation due to insufficient reference annotations.*

### Sequence Dropout Ablation

Sequence dropout experiments showed the model retained strong performance under input ablation (Figure 3). Mean WT Dice fell modestly from the full four-modality baseline (reported above) to 0.896 ± 0.006 across possible combinations of 3 inputs, 0.862 ± 0.014 for all possible combinations of 2 inputs, and 0.797 ± 0.022 across all single-modality inputs (mean ± SD across combinations). ET and TC followed a similar pattern; ED declined at a comparable relative rate, with its lower absolute performance reflecting the poor edema reference annotations described above.

**Figure 3:**
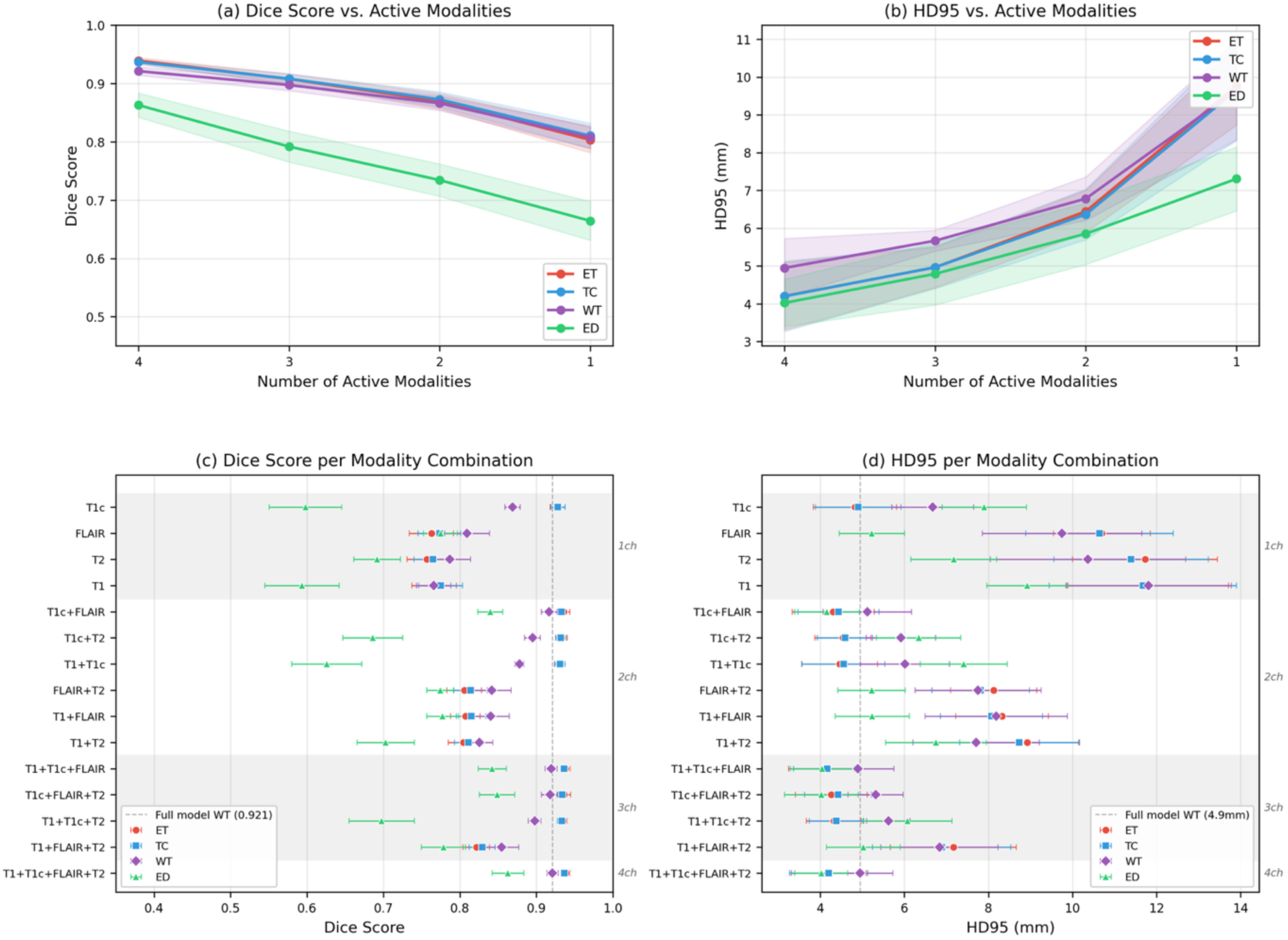
Sequence modality dropout ablation results. Figure showing segmentation performance for 4 tumor components after progressive removal of input MRI modalities. Metrics were computed per case on the held-out validation fold and pooled across all folds of the cross-validation. [A,B]Show mean Dice score (A) and 95th-percentile Hausdorff distance (HD95, mm) (B) performance over all possible combinations for each number of active input modalities All four components demonstrate graceful degradation; ED sits on a lower Dice baseline throughout, reflecting the poor edema reference annotations rather than a modality-dropout-specific effect. [C,D] Show per combination of Dice (C) and HD95 (D) for every possible subset of the four input modalities, grouped by number of active channels.

### External Validation Performance

In the external validation dataset, prior to exclusions, mean DSC and HD95 were (0.872 ± 0.118) and (10.708 ± 8.778 mm) for tumor core (n = 310), and (0.781 ± 0.175) and (12.512 ± 12.478 mm) for whole tumor (n = 310) (Table 3). Following edema exclusions, whole tumor metrics improved to (0.842 ± 0.117) and (7.255 ± 5.367 mm) for DSC and HD95 respectively (n = 156). Performance remained high despite heterogeneous imaging protocols and reduced modality completeness. Predicted tumor core volumes demonstrated strong correlation with reference volumes (r = 0.971, 95% CI [0.963, 0.977]; Figure 2C) but mild overestimation was observed for larger tumor volumes.

**Table 3:** Lesion-wise external validation (clinical dataset) metrics.

| Region | DSC |  | HD95 |  | *n* |
| --- | --- | --- | --- | --- | --- |
| | Mean ± Std | 95% CI | Mean ± Std | 95% CI | |
| TC | $0.872 \pm 0.118$ | [0.858, 0.885] | $10.708 \pm 8.778$ | [9.801, 11.783] | 310 |
| ED (before excl.) | $0.330 \pm 0.386$ | [0.287, 0.374] | $8.024 \pm 9.307$ | [6.552, 9.624] | 310 |
| ED (after excl.) | $0.673 \pm 0.270$ | [0.628, 0.715] | $8.024 \pm 9.307$ | [6.552, 9.624] | 156 |
| WT (before excl.) | $0.781 \pm 0.175$ | [0.761, 0.800] | $12.512 \pm 12.478$ | [11.191, 14.005] | 310 |
| WT (after excl.) | $0.842 \pm 0.117$ | [0.822, 0.859] | $7.255 \pm 5.367$ | [6.445, 8.177] | 156 |

*Note: TC= tumor core, ED = edema, WT = whole tumor. 154 cases did not have edema annotations and were not included in final edema and whole tumor evaluation. HD95 scores were computed over measurable overlap hence resulting in identical edema scores pre and post exclusion.*

### Failure Mode Analysis

Review of the lowest performing 5% of cases in the cross-validation cohort showed underperformance of tumors located in the skull base (n = 11), parafalcine tumors (n = 11), convexity tumors (n = 9), paraclinoid tumors (n = 6), posterior fossa tumors (n = 6) and other (n = 7). Differing failure modes were observed in the lowest 5% of clinical cases with underperformance primarily reported in intraosseous tumors (n = 11), stemming from poor scan quality (n = 2), and other (n = 3). During review, the model was observed to detect additional lesions in 9 cross-validation sample cases and 2 clinical sample cases.

Figure 4 demonstrates representative tumors with a spectrum of DSC values across each segmentation. The model demonstrated high sensitivity and identified several small convexity and parafalcine meningiomas that were absent from reference annotations, suggesting potential utility for lesion detection (Figure 4D).

**Figure 4:**
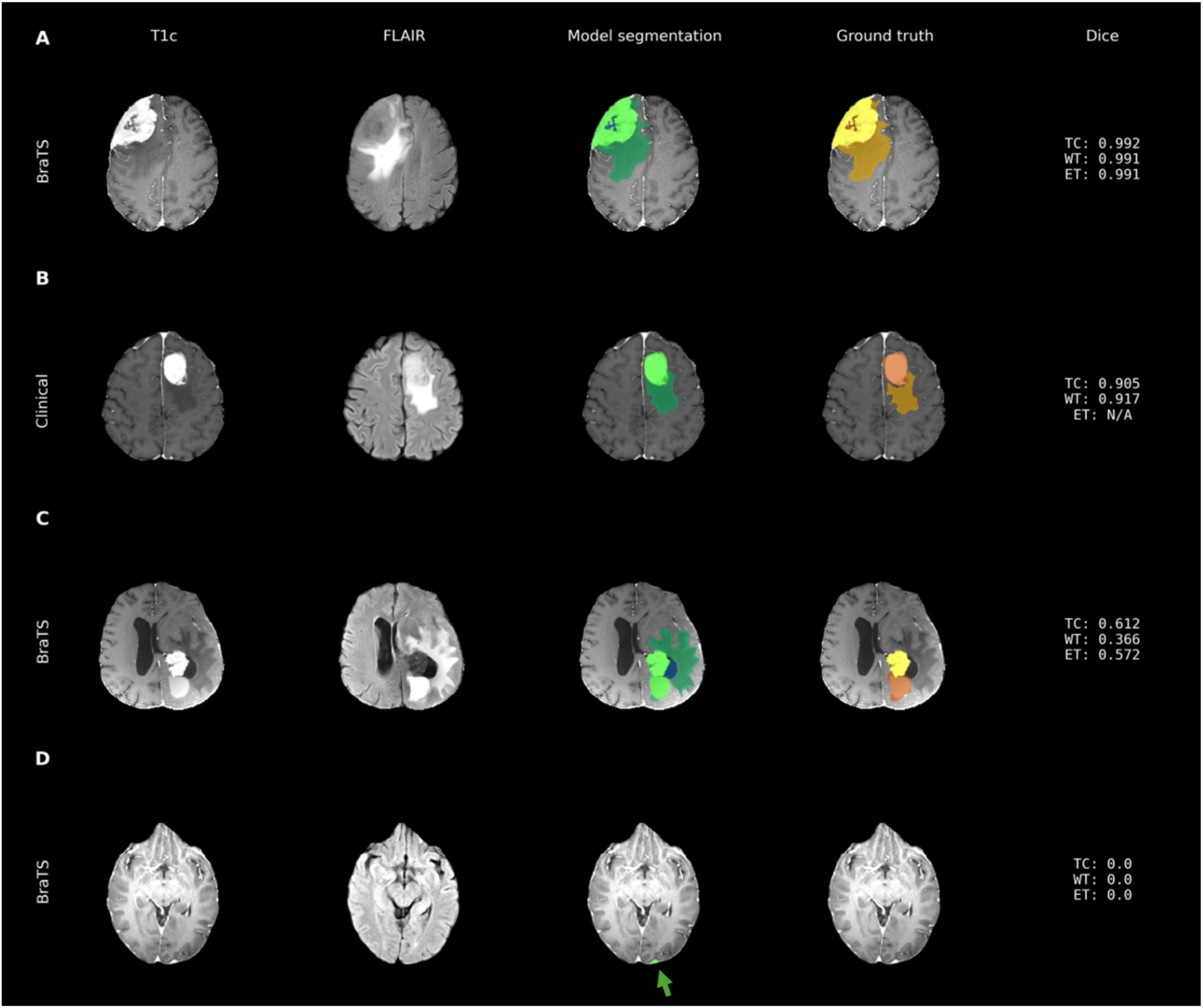
Representative tumor segmentations demonstrating the spectrum of DSC values in each compartment. Panel with representative segmentation results from the cross-validation (BraTS Meningioma 2023) and external validation (clinical) datasets showing variable concordance between nnU-Net predictions and expert annotations across diverse tumor locations and MR sequences. (A) demonstrates high concordance between model segmentation and reference annotations across all components. (B) demonstrates loss of concordance in TC and WT segmentations due to incomplete reference annotations. (C) demonstrates that the model mis-classifies T1W hyperintense tumor cyst as enhancing tumor but a separate tumor cyst is not included in reference TC resulting in reduced DSC for TC. (D) demonstrates a left convexity meningioma detected by the model that was absent from reference annotations.

### Model Performance Equity

Model performance was equitable across all three tumor components, with Gini indices of 0.052 for tumor core, 0.153 for edema, and 0.103 for whole tumor.

### Inference Performance

Inference required approximately 1.2 ± 0.9 seconds per case, compatible with integration into routine PACS workflows. A proposed PACS-integrated deployment pathway, returning DICOM segmentation objects and structured volumetric report fields for radiologist review, is illustrated in Figure 6.

**Figure 5:**
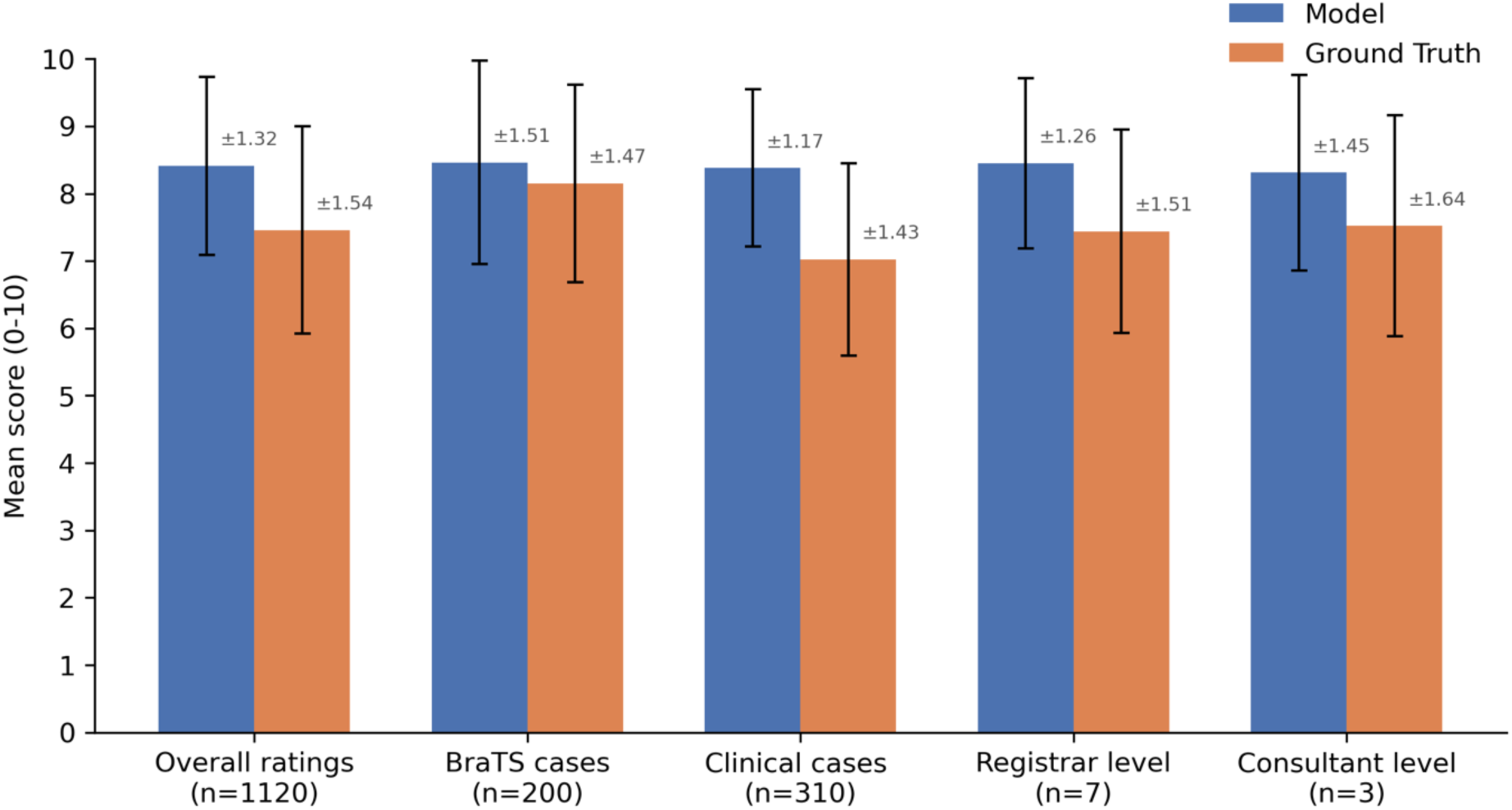
Clinical Rating of Model Segmentation versus Ground Truth Annotations. Bar chart showing the clinical ratings of the model segmentations in comparison to the corresponding reference annotations. The figure shows the model ratings outperforming reference annotations across different variables such as individual datasets and rater experience level.

**Figure 6:**
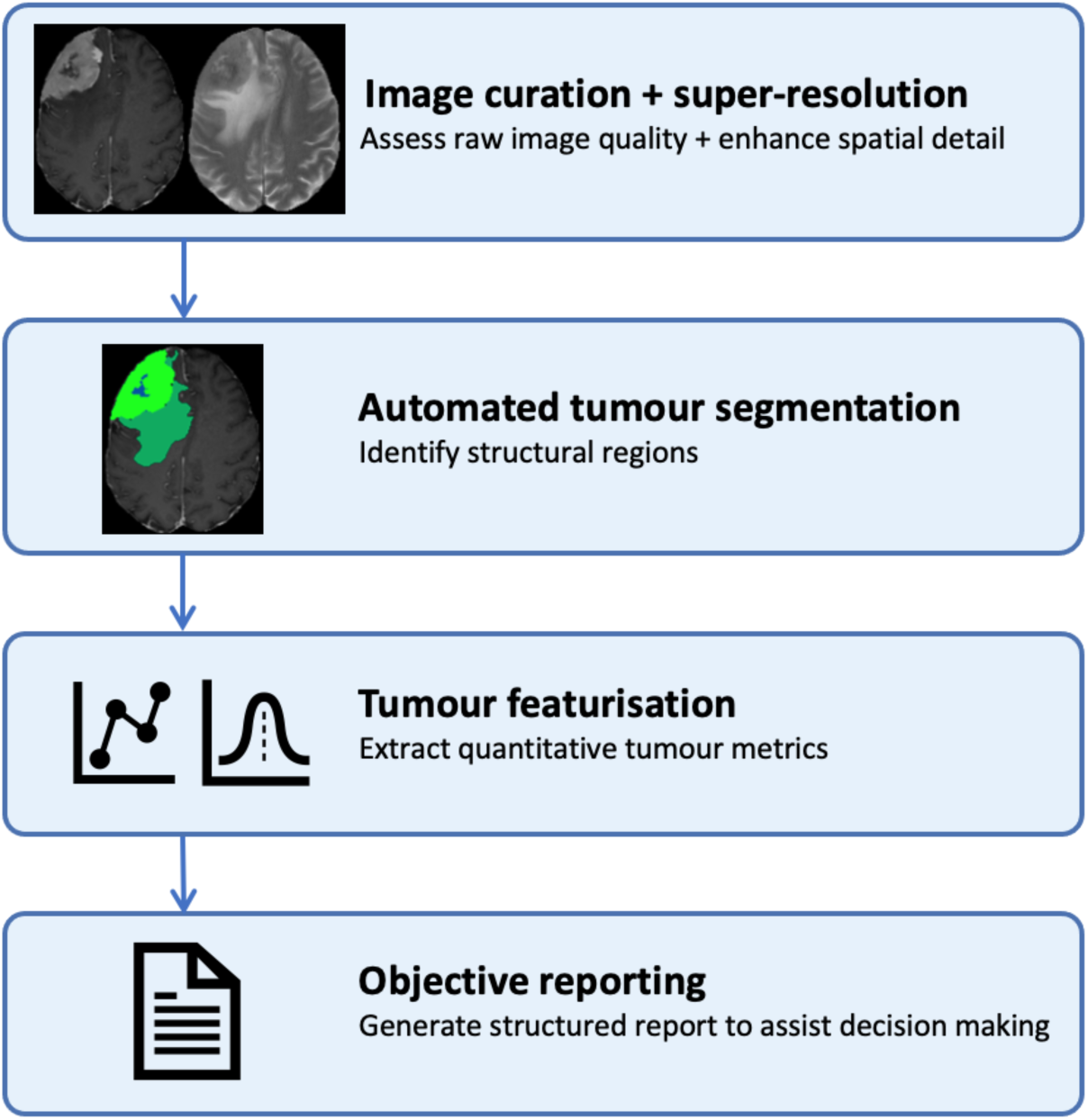
Potential radiology workflow integration flow chart. Workflow illustration demonstrating potential clinical integration of automated segmentation and featurization into radiology reporting and volumetric follow-up tools.

### Radiologist Ratings

A total of 1120 ratings were collected and analyzed from 10 raters across 510 cases, and a summary of results is reported in Figure 5. Inter-rater reliability was assessed on the subset of cases rated by at least 2 raters (50 cases, 100 paired observations). For reference annotation ratings, ICC for absolute agreement was 0.46 (95% CI [0.19, 0.58]) and consistency was 0.51 (95% CI [0.20, 0.64]); for model-generated segmentation ratings, both were 0.29 (95% CI [0.00, 0.42]). By published benchmarks (20) these values indicate poor to moderate reliability, consistent with the inherent subjectivity of segmentation quality rating tasks. The wide confidence intervals reflect that each overlap case was rated by approximately two raters, limiting ICC precision. Variance components from the LMM were SD = 0.639 for case, 0.258 for rater, 0.345 for rater × method, and 1.153 for residual error, indicating that case-level variability dominated rater-level variability (full random-effects table, Supplementary Table S2). Importantly, the LMM accounts for rater variability through random effects, meaning inter-rater disagreement does not invalidate the primary comparison; the ICC values characterize rating task difficulty rather than undermining the main result.

The mean quality score for model-generated segmentations was higher than for reference annotations. This pattern was observed across both datasets, with the difference being larger in the external validation dataset. A summary of results from the LMM model is shown in Table 4. Using the reference annotations as the reference category, the model-generated segmentations received numerically higher ratings (β = 0.32, 95% CI [−0.06, 0.69], p = 0.12); however, this overall effect was not statistically significant when taking into account rater × method variance. As described below, the model’s advantage was strongly dataset dependent.

**Table 4:**
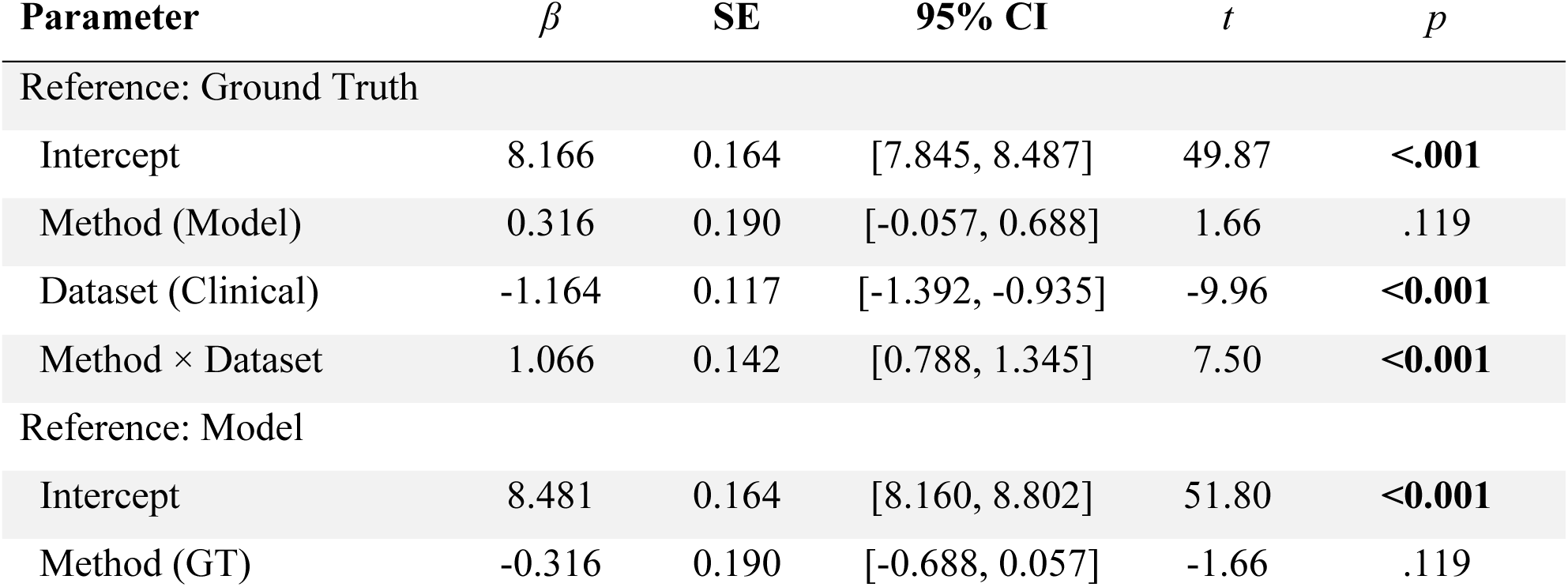
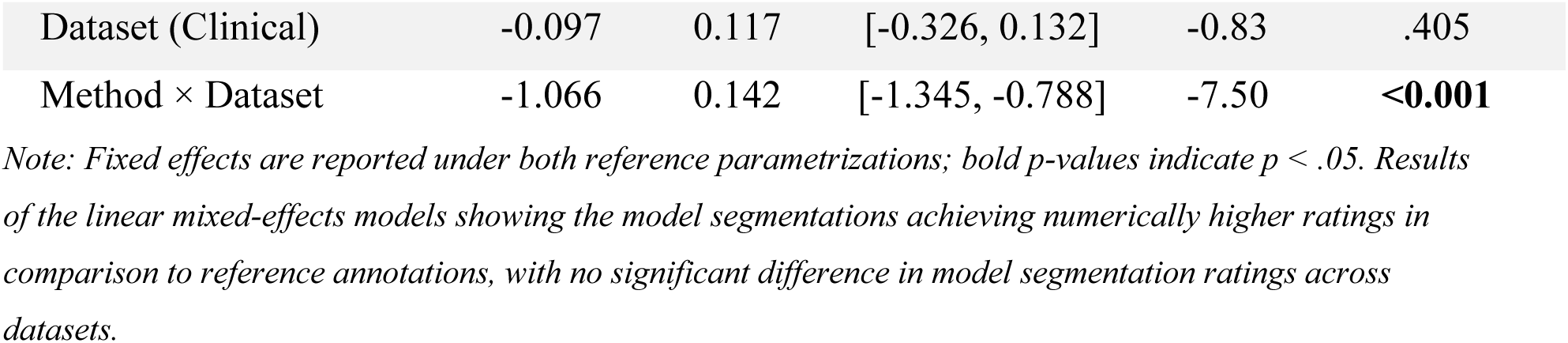
Fixed effects results of the linear mixed-effects models of rater scores.

| Parameter | $\beta$ | SE | 95% CI | *t* | *p* |
| --- | --- | --- | --- | --- | --- |
| Reference: Ground Truth |  |  |  |  |  |
| Intercept | 8.166 | 0.164 | [7.845, 8.487] | 49.87 | <.001 |
| Method (Model) | 0.316 | 0.190 | [-0.057, 0.688] | 1.66 | .119 |
| Dataset (Clinical) | -1.164 | 0.117 | [-1.392, -0.935] | -9.96 | <0.001 |
| Method × Dataset | 1.066 | 0.142 | [0.788, 1.345] | 7.50 | <0.001 |
| Reference: Model |  |  |  |  |  |
| Intercept | 8.481 | 0.164 | [8.160, 8.802] | 51.80 | <0.001 |
| Method (GT) | -0.316 | 0.190 | [-0.688, 0.057] | -1.66 | .119 |
| Dataset (Clinical) | -0.097 | 0.117 | [-0.326, 0.132] | -0.83 | .405 |
| Method × Dataset | -1.066 | 0.142 | [-1.345, -0.788] | -7.50 | **<0.001** |

*Note: Fixed effects are reported under both reference parametrizations; bold p-values indicate $p < .05$ . Results of the linear mixed-effects models showing the model segmentations achieving numerically higher ratings in comparison to reference annotations, with no significant difference in model segmentation ratings across datasets.*

A significant interaction effect between the method and dataset was observed, indicating that the model’s advantage over reference annotations was significantly larger in the external validation dataset compared with the BraTS Meningioma 2023 dataset. Specifically, the model scored 0.32 points higher than reference annotations in the BraTS Meningioma 2023 dataset, whereas this advantage increased to 1.38 points in the external validation dataset.

When the model was set as the reference category, the dataset effect became non-significant. This indicated that whereas reference annotation ratings differed significantly across datasets, the model-generated segmentations showed no significant difference in ratings across datasets indicating the model’s transferability to clinical scenarios. Furthermore, as the external validation dataset cases involved only one or two MRI sequences, this result provides further evidence suggesting the model is robust to channel dropout reinforcing the model’s adaptability.

## Discussion

A custom fully automated 3D nnU-Net residual encoder model achieved high segmentation accuracy for meningiomas in multi-institutional training data and generalized to an independent external validation dataset. In addition to overlap metrics, a blinded multi-rater study demonstrated higher radiologist-rated quality for model segmentations compared with reference annotations. Together, these findings support feasibility for integration into clinical workflows.

Prior meningioma segmentation studies have demonstrated feasibility using multiparametric MRI, but many relied on single-institution datasets with limited external validation; a recent meta-analysis highlighted variability in reported performance (11–14). Training on the large multi-institutional BraTS Meningioma 2023 dataset using a standardized self-configuring framework likely contributed to the generalizability observed here (10).

External validation demonstrated preserved performance in the external validation dataset with heterogeneous protocols. This is clinically important because follow-up examinations frequently include limited sequences. The use of augmentation and channel dropout was intended to improve robustness to missing modalities, which may contribute to performance in the external validation dataset.

Failure mode analysis identified skull base and intraosseous meningiomas as the most common sources of under-segmentation. These tumors have complex boundaries and limited contrast with adjacent bone and dura, and skull stripping may further degrade performance. Similar limitations have been described in prior work (13). Future work should focus on targeted enrichment of skull base and intraosseous cases and evaluation of skull-stripping–free pipelines.

Radiologists rated model segmentations numerically higher than reference annotations, particularly in the external validation dataset with the gap being four times larger in real-world clinical data than in the curated benchmark. This finding is consistent with known limitations of overlap metrics when reference annotations are imperfect (15,16). In clinical datasets, segmentation labels may be incomplete or inconsistent, particularly for edema. Therefore, radiologist-centered evaluation provides complementary evidence of clinical plausibility and may better reflect deployment suitability than DSC alone.

Automated meningioma segmentation may enable routine volumetry for surveillance imaging and post-treatment follow-up, addressing a longstanding gap in clinical workflows (6–8). The model is designed for server-based deployment, returning DICOM segmentation (DICOM SEG) objects to PACS within approximately 1.2 seconds of acquisition — well within same-session reporting time. The radiologist reviews and approves the segmentation before sign-off, preserving clinical accountability. Structured report fields would be populated automatically — for example: “Automated tumor core volume: 4.2 ml; change from prior (18 months): +0.6 ml (+16.7%; annualized growth rate: 0.4 ml/year)” — supporting standardized longitudinal documentation. Manual segmentation typically requires 20–40 minutes of specialist time per case; model inference requires less than two minutes including review, representing a workflow efficiency gain exceeding 90%. Automated segmentation further enables IBSI-compliant feature extraction across compartments — shape descriptors, enhancement heterogeneity, and peritumoral texture — supporting future radiomics applications including growth prediction and treatment response assessment (21,22).

Enhancing tumor performance could not be evaluated in the external validation dataset due to annotation constraints and edema overlays were excluded from radiologist evaluation because of inconsistent labeling. False-positive segmentation rates across other dural-based lesions were not assessed. Inter- and intra-rater variability of the reference segmentation annotations was not formally assessed; the ICC values reported relate to the Likert quality rating study rather than to annotation reproducibility. For the BraTS dataset, multi-reader consensus methodology is described in the source publication (17); however, annotation reliability for the single-institution external validation dataset was not formally quantified and represents a limitation of the reference standard. Methodologic guidance for clinical evaluation of artificial intelligence emphasizes the need for external validation, assessment of clinical effect, and prospective evaluation (23) which will be the focus of future work.

In conclusion, a fully automated 3D nnU-Net residual encoder model achieved high meningioma segmentation accuracy, generalized to independent clinical imaging with heterogeneous protocols, and produced radiologist-rated segmentations that exceeded reference annotation quality. Near-uniform performance across demographic subgroups, a sub-two-second inference time, and compatibility with PACS-integrated structured reporting together support feasibility for clinical deployment. Prospective evaluation is warranted before routine use for volumetric reporting and longitudinal surveillance in neuroradiology.

## Model availability

The model weights will be freely and openly available upon publication.

## Data Availability

The model weights will be freely and openly available upon publication.

