## Supplementary Material for "Deep Learning for Automated Meningioma Segmentation: Toward Clinical Integration and Workflow Efficiency"

*Fenney et al.*

### Supplementary Material

### Supplement S1. Linear Mixed-Effects Model Specification

#### Overview

Radiologist quality scores were analyzed using a linear mixed-effects model (LMM) implemented in R (version 4.3.2) using the *lme4* (version 1.1-35) and *lmerTest* (version 3.1-3) packages. Restricted maximum likelihood (REML) estimation was used throughout, with Satterthwaite approximation for degrees of freedom and *p*-values.

To account for the crossed structure of the experimental procedure, the results were analyzed using a linear mixed effects models (LMM) to compare segmentation quality scores between methods. The model was implemented in R using the lme4 and lmerTest packages with the following specification:

$$\text{Score} \sim \text{Method} * \text{Dataset} + (1|\text{Rater}) + (1|\text{Case}) + (1|\text{Rater:Method})$$

The model included fixed effects for method (model vs ground truth), dataset (BraTS Meningioma 2023 vs external validation), and their interaction. Random intercepts were included for rater and case to account for systematic differences such as scoring leniency and segmentation difficulty, and a rater × method random intercept captured rater-specific differences between the two methods. The model was fitted using restricted maximum likelihood (REML) estimation with Satterthwaite degrees of freedom for hypothesis testing. Inter-rater reliability was assessed separately on the subset of cases rated by at least 2 raters. A simpler intercept only mixed model defined as:

$$\text{Score} \sim 1 + (1|\text{Rater}) + (1|\text{Case})$$

was fit to this subset. ICC scores for both absolute agreement and consistency were estimated from the variance components of this simpler model.

#### Two Parameterisations

Results are reported under two parameterisations of the same model, differing only in which method level is set as the reference category:

**Parameterisation 1 — Reference annotations as reference category:** Positive coefficients for Method indicate model-generated segmentations scored higher than reference annotations. Used to report the primary method comparison and the interaction effect.

**Parameterisation 2 — Model-generated segmentations as reference category:** Allows direct testing of whether the model's ratings differed significantly across datasets (i.e. the dataset main effect under this parameterisation tests model transferability). A non-significant dataset effect under this parameterisation indicates the model performed consistently across BraTS and external validation data.

#### Inter-Rater Reliability Model

A separate intercept-only model was fitted to the subset of cases rated by at least two raters (50 cases; approximately 100 paired observations) to estimate inter-rater reliability:

```
Score ~ 1 + (1 | Rater) + (1 | Case)
```

Intraclass correlation coefficients (ICC) for absolute agreement and consistency were estimated from the variance components of this model. Let $\sigma^2 Rater$, $\sigma^2 Case$, and $\sigma^2 Residual$ denote the rater, case, and residual variance components respectively:

$$\text{ICC (absolute agreement)} = \sigma^2 Case \, / \, (\sigma^2 Case + \sigma^2 Rater + \sigma^2 Residual)$$

$$\text{ICC (consistency)} = \sigma^2 Case \, / \, (\sigma^2 Case + \sigma^2 Residual)$$

95% confidence intervals for ICC estimates were obtained by parametric bootstrap (1000 iterations). ICC values were interpreted using the benchmarks of Koo and Li (reference 20 in the main manuscript): <0.50 poor, 0.50–0.75 moderate, 0.75–0.90 good, >0.90 excellent. As noted in the manuscript, each overlap case was rated by approximately two raters; ICC estimates are therefore imprecise (wide 95% CIs) and should be interpreted as characterising rating task difficulty rather than as definitive reliability benchmarks.

#### Software and Reproducibility

All analyses were conducted in R version 4.3.2. Model fitting used `lme4::lmer()` with REML = TRUE. Hypothesis tests used `lmerTest::summary()` with Satterthwaite degrees of freedom. Fixed-effect estimates, standard errors, *t*-statistics, degrees of freedom, and *p*-values are reported in Table 4 of the main manuscript. Analysis code is available from the corresponding author on reasonable request.

---

#### Supplementary Table S1. Scanner Acquisition Parameters — External Validation Dataset

| Scanner | Field strength | Sequence | TR (ms) | TE (ms) | Flip angle (°) | Slice thickness (mm) | Matrix | Contrast dose (mmol/kg) |
|---|---|---|---|---|---|---|---|---|
| ***Siemens Avanto*** | 1.5T | T2 | 4420 | 90 | N/A | 5 | 512 x 464 | N/A |
| ***Siemens Avanto*** | 1.5T | FLAIR | 7000 | 300 | Variable: 120 to 50-70 | 1.1 | 461 x 512 | N/A |
| ***Siemens Avanto*** | 1.5T | T1W | 600 | 10 | Variable: 150 to 40-100 | 1 | 256 x 256 | N/A |
| ***Siemens Avanto*** | 1.5T | T1W+C | 600 | 10 | Variable 150 to 40-100 | 1 | 256 x 256 | 5 |
| ***Siemens Vida*** | 3T | T2 | 6400 | 80 | N/A | 4 | 640 x 640 | N/A |
| ***Siemens Vida*** | 3T | FLAIR | 7000 | 350 | Variable: 140 to 40-60 | 0.98 | 273 x 196 | N/A |
| ***Siemens Vida*** | 3T | T1W | 650 | 10 | Variable: 140 to 40-70 | 0.9 | 297 x 212 | N/A |
| ***Siemens Vida*** | 3T | T1W+C | 650 | 10 | Variable: 140 to 40-70 | 0.9 | 297 x 212 | 10 |

| | | | | | | | | |
|---|---|---|---|---|---|---|---|---|
| **Philips Achieva** | 3T | T2 | 3000 | 80 | N/A | 5 | 512 x 512 | N/A |
| **Philips Achieva** | 3T | FLAIR | 4800 | 350 | Variable:130 to 50-70 | 1 | 281 x 284 | N/A |
| **Philips Achieva** | 3T | T1W | 16.69 | 3 | 8-10 | 0.7 | 400 x 400 | N/A |
| **Philips Achieva** | 3T | T1W+C | 16.69 | 3 | 8-10 | 0.7 | 400 x 400 | 10 |

Abbreviations: TR, repetition time; TE, echo time; res., resolution; mmol/kg, millimoles per kilogram body weight. *Note:* Acquisition parameters for the BraTS Meningioma 2023 training dataset are described in the source publication (reference 17 in the main manuscript).

**Supplementary Table S2. Random effects results of the linear mixed-effects models of rater scores**

| **Random Effect** | **Variance** | **SD** |
|---|---|---|
| Rater | 0.067 | 0.258 |
| Case (stem) | 0.408 | 0.639 |
| Rater × Method | 0.119 | 0.345 |
| Residual | 1.328 | 1.153 |
